## Supplementary material for "Maternal obesity and metabolic disorders associate with congenital heart defects in the offspring: a systematic review": S1_Table

### Table S1: Search protocol

#### PubMed and Embase: Publications between January 1, 1990 and October 6, 2019.

##### Maternal overweight or obesity or metabolic syndrome and CHDs in offspring

*PubMed Search Strategy*

| | Aspect 1 | Aspect 2 | Aspect 3 |
|---|---|---|---|
| MeSH | Heart defects, congenital | Body Mass Index OR Overweight OR Adiposity OR Thinness OR Metabolic Syndrome | Pregnancy OR Infant OR Fetus |
| Free text search | Congenital heart defect* OR Congenital heart malformation* OR CHD OR Congenital heart disease* OR “Cardiac abnormalities” OR “Cardiac abnormality” OR Cardiac defect* OR Fetal heart OR Fetal cardiac defect* OR Foetal heart OR Foetal cardiac defect* | BMI OR Maternal overweight OR Maternal obesity OR Prepregnancy BMI OR Prepregnancy weight | Offspring* OR Pregnancy |

*PubMed Search 2019-10-06*

| | | No. of articles |
|---|---|---|
| #1 | "Heart Defects, Congenital"[Majr] | 121,469 |
| #2 | (((((((((Congenital heart defect*[Text Word]) OR Congenital heart malformation*[Text Word]) OR CHD[Text Word]) OR Congenital heart disease*[Text Word]) OR Cardiac abnormalities[Text Word]) OR Cardiac abnormality[Text Word]) OR Cardiac defect*[Text Word]) OR Fetal heart[Text Word]) OR Fetal cardiac defect*[Text Word]) OR Foetal heart[Text Word]) OR Foetal cardiac defect*[Text Word] | 105,996 |
| #3 | #1 OR #2<br>("Heart Defects, Congenital"[Majr]) OR ((((((((((Congenital heart defect*[Text Word]) OR Congenital heart malformation*[Text Word]) OR CHD[Text Word]) OR Congenital heart disease*[Text Word]) OR Cardiac abnormalities[Text Word]) OR Cardiac abnormality[Text Word]) OR Cardiac defect*[Text Word]) OR Fetal heart[Text Word]) OR Fetal cardiac defect*[Text Word]) OR Foetal heart[Text Word]) OR Foetal cardiac defect*[Text Word]) | 181,349 |
| #4 | (((("Body Mass Index"[Majr]) OR "Overweight"[Majr]) OR "Adiposity"[Majr]) OR "Thinness"[Majr]) OR "Metabolic Syndrome"[Majr] | 185,465 |
| #5 | (((Maternal overweight[Text Word]) OR Maternal obesity[Text Word]) OR Prepregnancy BMI[Text Word]) OR Prepregnancy weight[Text Word] | 3,677 |
| #6 | #4 OR #5<br>(((((("Body Mass Index"[Majr]) OR "Overweight"[Majr]) OR "Adiposity"[Majr]) OR "Thinness"[Majr]) OR "Metabolic Syndrome"[Majr])) OR ((((Maternal overweight[Text Word]) OR Maternal obesity[Text Word]) OR Prepregnancy BMI[Text Word]) OR Prepregnancy weight[Text Word]) | 187,082 |
| #7 | (("Pregnancy"[Majr]) OR "Infant"[Majr]) OR "Fetus"[Majr] | 273,767 |
| #8 | (Offspring*[Text Word]) OR Pregnancy[Text Word] | 975,184 |
| #9 | #7 OR #8<br>(((("Pregnancy"[Majr]) OR "Infant"[Majr]) OR "Fetus"[Majr])) OR ((Offspring*[Text Word]) OR Pregnancy[Text Word]) | 1,053,307 |
| #10 | #3 AND #6 AND #9<br>(((("Heart Defects, Congenital"[Majr]) OR ((((((((((Congenital heart defect*[Text Word]) OR Congenital heart malformation*[Text Word]) OR CHD[Text Word]) OR Congenital heart disease*[Text Word]) OR Cardiac abnormalities[Text Word]) OR Cardiac abnormality[Text Word]) OR Cardiac defect*[Text Word]) OR Fetal heart[Text Word]) OR Fetal cardiac defect*[Text Word]) OR Foetal heart[Text Word]) OR Foetal cardiac defect*[Text Word]))) AND ((((((("Body Mass Index"[Majr]) OR "Overweight"[Majr]) OR "Adiposity"[Majr]) OR "Thinness"[Majr]) OR "Metabolic Syndrome"[Majr])) OR ((((Maternal overweight[Text Word]) OR Maternal obesity[Text Word]) OR Prepregnancy BMI[Text Word]) OR Prepregnancy weight[Text Word]))) AND ((((("Pregnancy"[Majr]) OR "Infant"[Majr]) OR "Fetus"[Majr])) OR ((Offspring*[Text Word]) OR Pregnancy[Text Word])) | 114 |

*Embase Search Strategy*

| | Aspect 1 | Aspect 2 | Aspect 3 |
|---|---|---|---|

| | | | |
|---|---|---|---|
| ”MeSH” / Subject Heading<br><br>(Keyword + Map Term to Subject Heading + Explode + Include All Subheadings) | congenital heart malformation | exp body mass/ or exp obesity/ or exp underweight/<br><br>(“Obesity” broader term for “metabolic syndrome X”, “maternal obesity”, “morbid obesity”) | exp high risk pregnancy/ or exp pregnancy/ or infant/ or newborn/ or fetus/ |
| Free text search<br><br>(Keyword) | Congenital heart defect* OR Congenital heart malformation* OR CHD OR Congenital heart disease* OR Cardiac abnormalities OR Cardiac abnormality OR Cardiac defect* OR Fetal heart OR Fetal cardiac defect* OR Foetal heart OR Foetal cardiac defect* | Maternal overweight OR Maternal obesity OR Prepregnancy BMI OR Prepregnancy weight | Offspring* OR Pregnancy |

*Embase Search 2019-10-06*

| | | No. of articles |
|---|---|---|
| #1 | exp congenital heart malformation/ | 126,712 |
| #2 | Congenital heart defect* OR Congenital heart malformation* OR CHD OR Congenital heart disease* OR Cardiac abnormalities OR Cardiac abnormality OR Cardiac defect* OR Fetal heart OR Fetal cardiac defect* OR Foetal heart OR Foetal cardiac defect* | 128,917 |
| #3 | #1 OR #2 | 206,398 |
| #4 | exp high risk pregnancy/ or exp pregnancy/ or infant/ or newborn/ or fetus/ | 1,571,089 |
| #5 | Offspring* OR Pregnancy | 882,813 |
| #6 | #4 OR #5 | 1,768,205 |
| #7 | exp body mass/ or exp obesity/ or exp underweight/ | 761,522 |
| #8 | Maternal overweight OR Maternal obesity OR Prepregnancy BMI OR Prepregnancy weight | 7,737 |
| #9 | #7 OR #8 | 762,132 |
| #10 | #3 AND #6 AND #9 | 1,046 |
| LIMITS | English Language + Exclude MEDLINE journals | 91 |

#### Maternal diabetes and CHDs in offspring

PubMed Search Strategy

| | Aspect 1 | Aspect 2 | Aspect 3 |
|---|---|---|---|
| MeSH | Heart defects, congenital | Diabetes Mellitus OR Pregnancy in diabetics OR Hyperglycemia OR Insulin Resistance | Pregnancy OR Infant OR Fetus |
| Free text search | (as above) | Pregestational diabetes OR PGDM OR Maternal diabetes OR Gestational diabetes OR GDM OR Diabetic women OR Diabetic pregnancies | (as above) |

*PubMed Search 2019-10-06*

| | | No. of articles |
|---|---|---|
| #3 | #1 OR #2 (as above) | 181,349 |
| #4 | ((("Diabetes Mellitus"[Majr]) OR "Pregnancy in Diabetics"[Mesh]) OR "Hyperglycemia"[Majr]) OR "Insulin Resistance"[Majr] | 386,821 |
| #5 | ((((((Pregestational diabetes[Text Word]) OR PGDM[Text Word]) OR Maternal diabetes[Text Word]) OR Gestational diabetes[Text Word]) OR GDM[Text Word]) OR Diabetic women[Text Word]) OR Diabetic pregnancies[Text Word] | 20,405 |
| #6 | #4 OR #5<br>((((("Diabetes Mellitus"[Majr]) OR "Pregnancy in Diabetics"[Mesh]) OR "Hyperglycemia"[Majr]) OR "Insulin Resistance"[Majr])) OR (((((((Pregestational diabetes[Text Word]) OR PGDM[Text Word]) OR Maternal diabetes[Text | 393,455 |

| | | |
|---|---|---|
| | Word]) OR Gestational diabetes[Text Word]) OR GDM[Text Word]) OR Diabetic women[Text Word]) OR Diabetic pregnancies[Text Word]) | |
| #9 | #7 OR #8 (as above) | 1,053,307 |
| #10 | #3 AND #6 AND #9<br>(((((((("Diabetes Mellitus"[Majr]) OR "Pregnancy in Diabetics"[Mesh]) OR "Hyperglycemia"[Majr]) OR "Insulin Resistance"[Majr])) OR (((((((Pregestational diabetes[Text Word]) OR PGDM[Text Word]) OR Maternal diabetes[Text Word]) OR Gestational diabetes[Text Word]) OR GDM[Text Word]) OR Diabetic women[Text Word]) OR Diabetic pregnancies[Text Word]))) AND ((((("Pregnancy"[Majr]) OR "Infant"[Majr]) OR "Fetus"[Majr])) OR ((Offspring*[Text Word]) OR Pregnancy[Text Word]))) AND (("Heart Defects, Congenital"[Majr]) OR ((((((((((Congenital heart defect*[Text Word]) OR Congenital heart malformation*[Text Word]) OR CHD[Text Word]) OR Congenital heart disease*[Text Word]) OR Cardiac abnormalities[Text Word]) OR Cardiac abnormality[Text Word]) OR Cardiac defect*[Text Word]) OR Fetal heart[Text Word]) OR Fetal cardiac defect*[Text Word]) OR Foetal heart[Text Word]) OR Foetal cardiac defect*[Text Word])) | 632 |

Embase Search Strategy

| | Aspect 1 | Aspect 2 | Aspect 3 |
|---|---|---|---|
| "MeSH" / Subject Heading | congenital heart malformation | exp diabetes mellitus/ or exp hyperglycemia/ or exp insulin resistance/ | exp high risk pregnancy/ or exp pregnancy/ or infant/ or newborn/ or fetus/ |
| Free text search | (as above) | Pregestational diabetes OR PGDM OR Maternal diabetes OR Gestational diabetes OR GDM OR Diabetic women OR Diabetic pregnancies | (as above) |

*Embase Search 2019-10-06*

| | | No. of articles |
|---|---|---|
| #3 | #1 OR #2 (as above) | 206,398 |
| #6 | #4 OR #5 (as above) | 1,768,205 |
| #7 | exp diabetes mellitus/ or exp hyperglycemia/ or exp insulin resistance/ | 1,005,513 |
| #8 | Pregestational diabetes OR PGDM OR Maternal diabetes OR Gestational diabetes OR GDM OR Diabetic women OR Diabetic pregnancies | 32,162 |
| #9 | #7 OR #8 | 1,008,162 |
| #10 | #3 AND #6 AND #9 | 2,001 |
| LIMITS | English Language + Exclude MEDLINE journals | 182 |

#### Maternal hypertension or preeclampsia and CHDs in offspring

*PubMed Search Strategy*

| | Aspect 1 | Aspect 2 | Aspect 3 |
|---|---|---|---|
| MeSH | Heart defects, congenital | Hypertension OR Hypertension, pregnancy-induced | Pregnancy OR Infant OR Fetus |
| Free text search | (as above) | Maternal hypertension OR Maternal hypertensive disorder* OR Gestational hypertension | (as above) |

*PubMed Search 2019-10-06*

| | | No. of articles |
|---|---|---|
| #3 | #1 OR #2 (as above) | 181,349 |
| #4 | ("Hypertension"[Majr]) OR "Hypertension, Pregnancy-Induced"[Mesh] | 211,800 |
| #5 | ((Maternal hypertension[Text Word]) OR Maternal hypertensive disorder*[Text Word]) OR Gestational hypertension[Text Word] | 3,466 |
| #6 | #4 OR #5<br>((("Hypertension"[Majr]) OR "Hypertension, Pregnancy-Induced"[Mesh])) OR (((Maternal hypertension[Text Word]) OR Maternal hypertensive disorder*[Text Word]) OR Gestational hypertension[Text Word]) | 213,166 |
| #9 | #7 OR #8 (as above) | 1,053,307 |

| | | |
|---|---|---|
| #10 | #3 AND #6 AND #9<br>((((("Hypertension"[Majr]) OR "Hypertension, Pregnancy-Induced"[Mesh])) OR (((Maternal hypertension[Text Word]) OR Maternal hypertensive disorder*[Text Word]) OR Gestational hypertension[Text Word]))) AND (((("Pregnancy"[Majr]) OR "Infant"[Majr]) OR "Fetus"[Majr])) OR ((Offspring*[Text Word]) OR Pregnancy[Text Word]))) AND (("Heart Defects, Congenital"[Majr]) OR (((((((((Congenital heart defect*[Text Word]) OR Congenital heart malformation*[Text Word]) OR CHD[Text Word]) OR Congenital heart disease*[Text Word]) OR Cardiac abnormalities[Text Word]) OR Cardiac abnormality[Text Word]) OR Cardiac defect*[Text Word]) OR Fetal heart[Text Word]) OR Fetal cardiac defect*[Text Word]) OR Foetal heart[Text Word]) OR Foetal cardiac defect*[Text Word])) | 661 |

*Embase Search Strategy*

| | Aspect 1 | Aspect 2 | Aspect 3 |
|---|---|---|---|
| ”MeSH” / Subject Heading | congenital heart malformation | exp hypertension/ or exp "eclampsia and preeclampsia"/ | exp high risk pregnancy/ or exp pregnancy/ or infant/ or newborn/ or fetus/ |
| Free text search | (as above) | Maternal hypertension OR Maternal hypertensive disorder* OR Gestational hypertension | (as above) |

*Embase Search 2019-10-06*

| | | No. of articles |
|---|---|---|
| #3 | #1 OR #2 (as above) | 206,398 |
| #6 | #4 OR #5 (as above) | 1,768,205 |
| #7 | exp hypertension/ or exp "eclampsia and preeclampsia"/ | 735,885 |
| #8 | Maternal hypertension OR Maternal hypertensive disorder* OR Gestational hypertension | 18,442 |
| #9 | #7 OR #8 | 736,285 |
| #10 | #3 AND #6 AND #9 | 2,637 |
| LIMITS | English Language + Exclude MEDLINE journals | 270 |

#### Maternal dyslipidemia and CHDs in offspring

*PubMed Search Strategy*

| | Aspect 1 | Aspect 2 | Aspect 3 |
|---|---|---|---|
| MeSH | Heart defects, congenital | Dyslipidemias OR Lipoproteins, HDL | Pregnancy OR Infant OR Fetus |
| Free text search | (as above) | HDL OR Dyslipidemia* | (as above) |

*PubMed Search 2019-10-06*

| | | No. of articles |
|---|---|---|
| #3 | #1 OR #2 (as above) | 181,349 |
| #4 | ("Dyslipidemias"[Majr]) OR "Lipoproteins, HDL"[Majr] | 66,228 |
| #5 | (Dyslipidemia*[Text Word]) OR HDL[Text Word] | 103,854 |
| #6 | #4 OR #5<br>((("Dyslipidemias"[Majr]) OR "Lipoproteins, HDL"[Majr])) OR ((Dyslipidemia*[Text Word]) OR HDL[Text Word]) | 138,647 |
| #9 | #7 OR #8 (as above) | 1,053,307 |
| #10 | #3 AND #6 AND #9<br>(#52 AND #96 AND #62)<br>(((((("Dyslipidemias"[Majr]) OR "Lipoproteins, HDL"[Majr])) OR ((Dyslipidemia*[Text Word]) OR HDL[Text Word]))) AND (((("Pregnancy"[Majr]) OR "Infant"[Majr]) OR "Fetus"[Majr])) OR ((Offspring*[Text Word]) OR Pregnancy[Text Word]))) AND (("Heart Defects, Congenital"[Majr]) OR ((((((((((Congenital heart defect*[Text Word]) OR Congenital heart malformation*[Text Word]) OR CHD[Text Word]) OR Congenital heart disease*[Text Word]) OR Cardiac abnormalities[Text Word]) OR Cardiac abnormality[Text Word]) OR Cardiac defect*[Text Word]) OR Fetal heart[Text Word]) OR Fetal cardiac defect*[Text Word]) OR Foetal heart[Text Word]) OR Foetal cardiac defect*[Text Word])) | 87 |

*Embase Search Strategy*

| | Aspect 1 | Aspect 2 | Aspect 3 |
|---|---|---|---|

| | | | |
|---|---|---|---|
| ”MeSH” / Subject Heading | congenital heart malformation | exp dyslipidemia/ or exp hypertriglyceridemia/ or exp high density lipoprotein/ | exp high risk pregnancy/ or exp pregnancy/ or infant/ or newborn/ or fetus/ |
| Free text search | (as above) | HDL OR Dyslipidemia* | (as above) |

*Embase Search 2019-10-06*

| | | No. of articles |
|---|---|---|
| #3 | #1 OR #2 (as above) | 206,398 |
| #6 | #4 OR #5 (as above) | 1,768,205 |
| #7 | exp dyslipidemia/ or exp hypertriglyceridemia/ or exp high density lipoprotein/ | 145,111 |
| #8 | HDL OR Dyslipidemia* | 166,506 |
| #9 | #7 OR #8 | 202,390 |
| #10 | #3 AND #6 AND #9 | 187 |
| LIMITS | English Language + Exclude MEDLINE journals | 17 |
