## Supplementary material for "Maternal obesity and metabolic disorders associate with congenital heart defects in the offspring: a systematic review": S2_Table

Table S2: Newcastle-Ottawa Quality Assessment Scale – Case-control studies

| Selection | | |
|---|---|---|
| 1. Is the case definition adequate? | A. yes, with independent validation | ☆ |
| | B. yes, eg record linkage or based on self-reports | |
| | C. no description | |
| 2. Representativeness of the cases | A. consecutive or obviously representative series of cases | ☆ |
| | B. potential for selection biases or not stated | |
| 3. Selection of Controls | A. community controls | ☆ |
| | B. hospital controls | |
| | C. no description | |
| 4. Definition of Controls | A. no history of disease (endpoint) | ☆ |
| | B. no description of source | |
| Comparability | | |
| 1. Comparability of cases and controls on the basis of the design or analysis | A. study controls for ________________ (Select the most important factor.) | ☆ |
| | B. study controls for any additional factor (This criteria could be modified to indicate specific control for a second important factor.) | ☆ |
| Exposure | | |
| 1. Ascertainment of exposure | A. secure record (eg surgical records) | ☆ |
| | B. structured interview where blind to case/control status | ☆ |
| | C. interview not blinded to case/control status | |
| | D. written self-report or medical record only | |
| | E. no description | |
| 2. Same method of ascertainment for cases and controls | A. yes | ☆ |
| | B. no | |
| 3. Non-Response rate | A. same rate for both groups | ☆ |
| | B. non respondents described | |
| | C. rate different and no designation | |

Note: A study can be awarded a maximum of one star for each numbered item within the Selection and Exposure categories. A maximum of two stars can be given for Comparability.

Further explanations and manuals for the scale can be found at:
http://www.ohri.ca/programs/clinical_epidemiology/oxford.asp

Table S2 (continued): Newcastle-Ottawa Quality Assessment Scale – Cohort studies

| | | |
|---|---|---|
| Selection | | |
| 1. Representativeness of the exposed cohort | A. truly representative of the average ________________ (describe) in the community | ☆ |
| | B. somewhat representative of the average _______________ in the community | ☆ |
| | C. selected group of users eg nurses, volunteers | |
| | D. no description of the derivation of the cohort | |
| 2. Selection of the non-exposed cohort | A. drawn from the same community as the exposed cohort | ☆ |
| | B. drawn from a different source | |
| | C. no description of the derivation of the non-exposed cohort | |
| 3. Ascertainment of exposure | A. secure record (eg surgical records) | ☆ |
| | B. structured interview | ☆ |
| | C. written self-report | |
| | D. no description | |
| 4. Demonstration that outcome of interest was not present at start of study | A. yes | ☆ |
| | B. no | |
| Comparability | | |
| 1. Comparability of cohorts on the basis of the design or analysis | A. study controls for ________________ (Select the most important factor.) | ☆ |
| | B. study controls for any additional factor (This criteria could be modified to indicate specific control for a second important factor.) | ☆ |
| Outcome | | |
| 1. Assessment of outcome | A. independent blind assessment | ☆ |
| | B. record linkage | ☆ |
| | C. self-report | |
| | D. no description | |
| 2. Was follow-up long enough for outcomes to occur | A. yes (select an adequate follow up period for outcome of interest) | ☆ |
| | B. no | |
| 3. Adequacy of follow up of cohorts | A. complete follow up - all subjects accounted for | ☆ |
| | B. subjects lost to follow up unlikely to introduce bias - small number lost - > ____ % (select an adequate %) follow up, or description provided of those lost) | ☆ |
| | C. follow up rate < ____% (select an adequate %) and no description of those lost | |
| | D. no statement | |

Note: A study can be awarded a maximum of one star for each numbered item within the Selection and Outcome categories. A maximum of two stars can be given for Comparability.

Further explanations and manuals for the scale can be found at:
http://www.ohri.ca/programs/clinical_epidemiology/oxford.asp
