## Supplementary material for "Maternal obesity and metabolic disorders associate with congenital heart defects in the offspring: a systematic review": S2_Table

Table S3: Characteristics of included studies

| Author/publication year | Country of study population | Study design | Population size | Cases with CHDs | Included study population | Plurality | Maternal exposure | Sources of maternal exposure | NOS score |
|---|---|---|---|---|---|---|---|---|---|
| Hoang 2016 [17] | USA (Texas) | Cohort | 4 207 898 | 48 249 | Live births | All | PGDM (not specified), GDM | Registers | 8 |
| Persson 2019 [14] | Sweden | Cohort | 2 050 491 | 28 628 | Live births | Singletons | Obesity | Registers | 8 |
| Liu 2013 [19] | Canada (excl. Quebec) | Cohort | 2 278 838 | 26 488 | Live births | All | Obesity, DM1, DM2, hypertension | Registers | 8 |
| Chou 2016 [46] | Taiwan | Cohort | 1 387 650 | 23 483 | Live births | All | DM1, DM2, hypertension | Registers | 8 |
| Boyd 2017 [21] | Denmark | Cohort | 1 972 857 | 18 038 | Live births | Singletons | PE | Registers | 8 |
| Auger 2015 [22] | Canada (Quebec) | Cohort | 1 942 072 | 17 296 | Live births | All | PE | Registers | 8 |
| Ludvigsson 2018 [13] | Sweden | Cohort | 1 162 323 | 17 242 | Live births | Singletons | DM1 | Registers | 8 |
| Øyen 2016 [16] | Denmark | Cohort | 2 025 727 | 16 325 | Live births | Singletons | PGDM (DM1 or DM2), GDM | Registers | 8 |
| Blomberg 2010 [30] | Sweden | Cohort | 1 049 582 | 11 163 | Live births, stillbirths | Not stated | Obesity | Registers | 8 |
| Fisher 2017 [20] | USA | Case-control (NBDPS) | 21 762 | 10 625 | Live births, stillbirths, terminated | Singletons | Untreated hypertension | Interviews | 8 |
| Leirgul 2016 [18] | Norway | Cohort | 914 427 | 10 575 | Live births, stillbirths, terminated | Singletons | PGDM (DM1, DM2 or unspecified DM), GDM | Registers | 8 |
| Block 2013 [29] | USA (Florida) | Case-control | 1 124 370 | 9 314 | Live births | Singletons | Obesity | Registers | 7 |
| Mills 2010 [32] | USA (New York State) | Case-control (nested) | 63 696 | 7 392 | Live births | Singletons | Obesity | Medical records | 7 |
| Cedergren 2003 [35] | Sweden | Case-control | 812 457 | 6 801 | Live births, stillbirths | Not stated | Obesity | Registers | 7 |
| Gilboa 2010 [31] | USA | Case-control (NBDPS) | 12 113 | 6 440 | Live births, stillbirths | Not stated | Obesity, obesity+GDM | Interviews | 8 |
| Correa 2008 [44] | USA | Case-control (NBDPS) | 17 925 | 4 621 | Live births | Not stated | PGDM (DM1 or DM2), GDM | Interviews | 8 |
| Vereczkey 2014 [41] | Hungary | Case-control | 41 713 | 3 562 | Live births | All | PGDM (DM1 or DM2), hypertension | Registers | 8 |
| Brodwall 2016 [48] | Norway | Cohort | 914 703 | 2 473 | Live births, stillbirths, terminated | Singletons | PE | Registers | 8 |

| | | | | | | | | | |
|---|---|---|---|---|---|---|---|---|---|
| Sharpe 2005 [47] | Australia (South Australia) | Cohort | 282 260 | 2 418 | Live births, stillbirths | Singletons | DM1, GDM (GDM or impaired glucose tolerance) | Interviews | 8 |
| Liu 2015 [39] | China (Tianjin) | Cohort | 90 796 | 1 817 | Live births | All | Obesity, GDM | Questionnaire / medical records | 8 |
| Brite 2014 [15] | USA | Cohort (CSL) | 121 815 | 1 388 | Live births | Singletons | Obesity | Medical records | 8 |
| Watkins 2001 [37] | USA | Case-control (ABDCCS) | 4 078 | 1 049 | Live births, stillbirths | Not stated | Obesity | Interviews | 7 |
| Agopian 2012 [40] | USA (Texas) | Cohort | 3 806 299 | 563 | Live births, stillbirths, terminated | All | Obesity, PGDM (not specified), GDM | Registers | 8 |
| Rankin 2010 [33] | England | Cohort | 41 013 | 341 | Live births, stillbirths, terminated | Singletons | Obesity | Registers | 8 |
| Shaw 2008 [34] | USA (California) | Case-control | 1 359 | 323 | Live births, stillbirths, terminated | Not stated | Obesity | Interviews | 8 |
| Kovalenko 2018 [42] | Russia (Murmansk) | Cohort | 52 253 | 233 | Live births | Singletons | PGDM (DM1 or DM2) | Registers | 8 |
| Shaw 2000 [38] | USA (California) | Case-control | 2 033 | 202 | Live births, stillbirths | Not stated | Obesity | Interviews | 7 |
| Watkins 2003 [36] | USA | Case-control (Atlanta BDRFSS) | 525 | 195 | Live births, stillbirths, terminated | Not stated | Obesity | Interviews | 7 |
| Vinceti 2014 [43] | Italy | Case-control (nested) | 479 720 | 68 | Live births, stillbirths | All | PGDM (DM1 or DM2) | Registers | 8 |
| Erickson 1991 [45] | USA | Case-control (ABDCCS) | 7 900 | 35 | Live births, stillbirths | Not stated | PGDM (not specified) | Interviews | 7 |

Note: Studies listed after number of CHD cases: studies with most cases first etc.

Abbreviations: ABDCCS, Atlanta Birth Defect Case-Control Study; BDRFSS, Birth Defects Risk Factor Surveillance Study; CHDs, congenital heart defects; CM, congenital malformations; CSL, The Consortium of Safe Labor; DM1, diabetes mellitus type 1; DM2, diabetes mellitus type 2; GDM, gestational diabetes mellitus; NBDPS, National Birth Defects Prevention Study; NOS, Newcastle Ottawa Scale; PE, preeclampsia; PGDM, pregestational diabetes.
